# Specialty-type and state-level variation in paroxetine use among older adult patients

**DOI:** 10.1101/2023.02.15.23285973

**Authors:** Luke R. Cavanah, Jessica L. Goldhirsh, Leighton Y. Huey, Brian J. Piper

## Abstract

**Introduction:** Paroxetine is an older “selective” serotonin reuptake inhibitor (SSRI) that is notable for its lack of selectivity, resulting in a cholinergic adverse-effect profile, especially among older adults (65+).

**Methods:** Paroxetine prescription rates and costs per state were ascertained from the Medicare Specialty Utilization and Payment Data. States’ annual prescription rate, corrected per thousand Part D enrollees, outside 95% confidence interval were considered significantly different from the average.

**Results:** There was a steady decrease in paroxetine prescriptions (-34.52%) and spending (-16.69%) from 2015-2020 but a consistent, five-fold state-level difference. From 2015-2020, Kentucky (194.9, 195.3, 182.7, 165.1, 143.3, 132.5) showed significantly higher prescriptions rates relative to the national average, and Hawaii (42.1, 37.9, 34.3, 31.7, 27.7, 26.6) showed significantly lower prescription rates. North Dakota was often a frequent elevated prescriber of paroxetine (2016: 170.7, 2018: 143.3), relative to the average. Neuropsychiatry and geriatric medicine frequently prescribed the largest amount of paroxetine prescriptions, relative to the number of providers in that specialty, from 2015-2020.

**Discussion:** Despite the American Geriatrics Society prohibition against paroxetine use in the older adults and many effective treatment alternatives, paroxetine was still commonly used in this population, especially in Kentucky and North Dakota and by neuropsychiatry and geriatric medicine. These findings provide information on the specialty types and states where education and policy reform would likely have the greatest impact on improving adherence to the paroxetine prescription recommendations.

## Introduction

Despite their name, “selective” serotonin reuptake inhibitors (SSRIs) often have activity on other chemicals, which can contribute to adverse effects (Ferguson, 2001; Stahl, 2021). One SSRI that is particularly infamous for its nonselective action on other neurotransmitters and liver enzymes is paroxetine (Nevels et al., 2016). Paroxetine was first marketed in the US in 1992, and it is indicated for use in depression, obsessive compulsive disorder, panic disorder, social anxiety disorder, and generalized anxiety disorder. Paroxetine is available in generic and brand names (Paxil ®, Pexeva ®, Brisdelle®). Figure 1 illustrates that in addition to its notable target at the serotonin reuptake transporter (SERT), it has appreciable affinity for the norepinephrine transporter (NET), central muscarinic (M_1_) receptors, nitric oxide synthase (NOS) (Finkel et al., 1996), and CYP 2D6 (Schatzberg & Nemeroff, 2017; Stahl, 2021). Some research also suggests paroxetine additionally has activity at CYP 3A4 (Zanger & Schwab, 2013). As a consequence of the lack of specificity, paroxetine, compared to other SSRIs, results in increased sedation, constipation, sexual dysfunction, discontinuation syndrome and weight gain (Marks et al., 2008).

**Figure 1.**
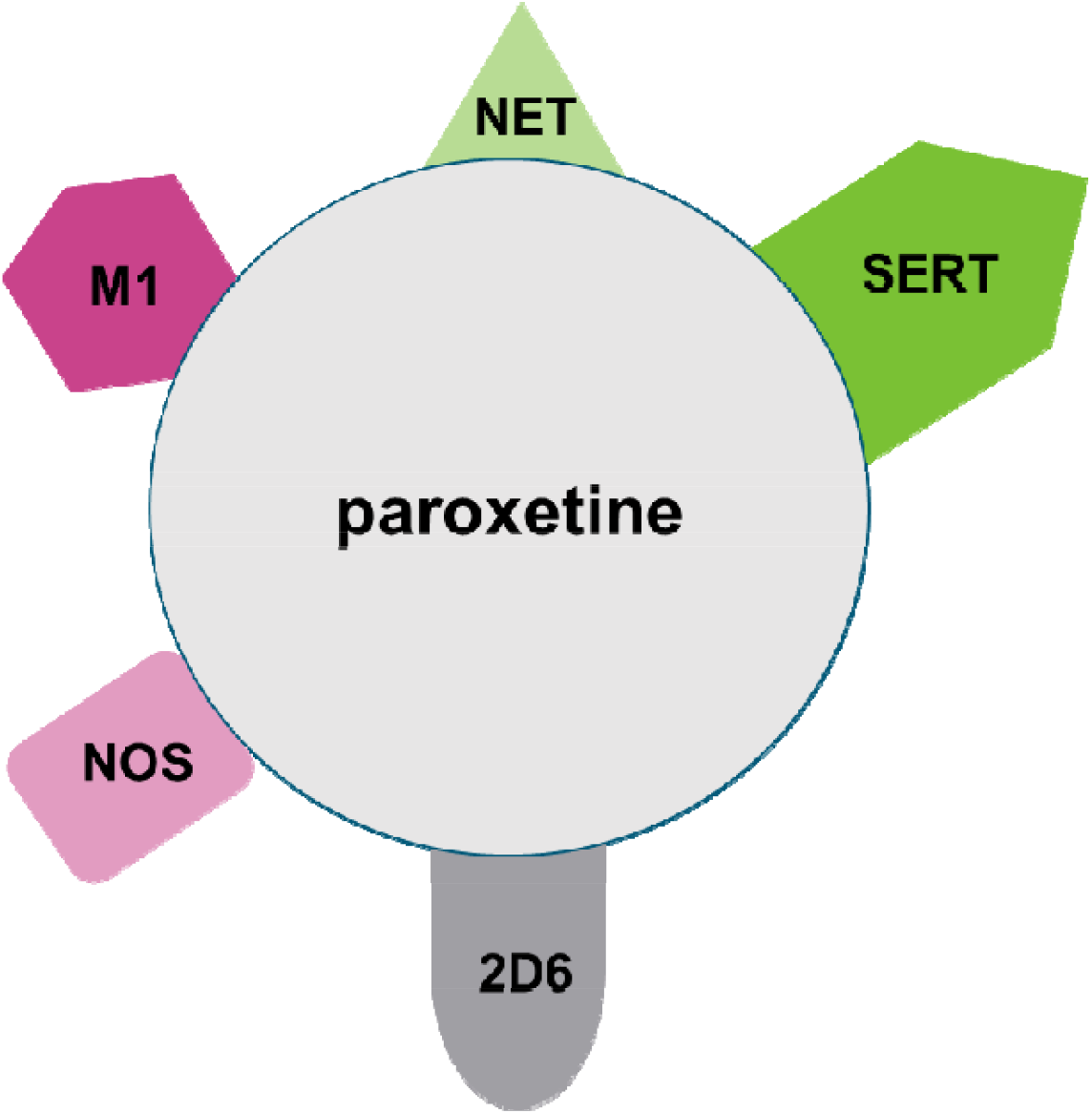
Mechanisms of action of paroxetine. Paroxetine has appreciable affinity for serotonin reuptake transporter (SERT), norepinephrine transporter (NET), central muscarinic (M_1_) receptors, nitric oxide synthase (NOS), and CYP 2D6.

Compared to all SSRI medications, paroxetine has the highest affinity for the M_1_ receptor (*K*_*i*_ = 76 nM). Consequently, paroxetine use is associated with the classical anticholinergic side effects, such as constipation, urinary retention, increased intraocular pressure, blurred vision, dry mouth, dry eyes, flushing, and hyperthermia (Jacob & Spinler, 2006; Julien, 2013; Schatzberg & Nemeroff, 2017; Stahl, 2020). Older adult populations are particularly at risk to anticholinergic effects, so use of anticholinergic medications, such as paroxetine, in the older adults should be reduced when clinically appropriate, especially in those suffering from dementia (Kalisch Ellett et al., 2014; Sakakibara et al., 2008; Tune, 2001). Furthermore, anticholinergic drugs have been demonstrated to contribute to cognitive decline and dementia in older adults (Carrière et al., 2009). Syndrome of Inappropriate Anti-Diuretic Hormone (SIADH) is also a well-known, concerning adverse effect of various SSRIs, and older age is a risk factor of SSRI-induced SIADH due to age-related changed in renal functioning (Jacob & Spinler, 2006; Kirby et al., 2002).

In addition to paroxetine’s lack of receptor selectivity with appreciable affinity for the norepinephrine transporter and nitric oxide (Finkel et al., 1996), it also has many medication interactions due to action on the CYP450 superfamily (Nevels et al., 2016). Paroxetine is a strong inhibitor of P450 3A4 isoenzyme, which metabolizes approximately 50% of prescribed drugs (Zanger & Schwab, 2013). Moreover, paroxetine is the strongest inhibitor of P450 2D6 isoenzyme (*K*_*i*_ = 0.065-4.65 μM) of all antidepressants (Schatzberg & Nemeroff, 2017; Stahl, 2021). CYP2D metabolizes many medications, such as antipsychotics, tricyclic antidepressants, class IC antiarrhythmics, β-adrenergic agents, trazodone, and dextromethorphan (Nemeroff et al., 1996). Older adults tend to take more medications, with an average of six to eight (Buck et al., 2009; D.-C. Chan et al., 2012; Chrischilles et al., 2007), increasing the risk of drug-drug interactions.

Adverse drug events in older adults are certainly not unique to paroxetine: approximately 15% of hospitalizations of this population are secondary to an adverse drug event (Beijer & de Blaey, 2002); this value is double for those over 75 (M. Chan et al., 2001; Page & Ruscin, 2006). In the outpatient setting, it is estimated that approximately 30% of older adults are taking potentially inappropriate medications (PIMs) (Insani et al., 2021). Approximately 30-50% of adverse drug events are preventable (M. Chan et al., 2001; Gurwitz et al., 2003). Clearly, minimizing such adverse events has significant health and economic consequences on both a micro and a macro scale.

Leading professional organizations in the field of geriatrics are clear that paroxetine is a potentially higher risk medication that should be avoided in older adults when possible (American Geriatrics Society Beers Criteria® Update Expert Panel, 2019). There is high quality evidence indicating that paroxetine is strongly anticholinergic and has an unfavorable likelihood of causing sedation and orthostatic hypertension and thus, it is strongly recommended that paroxetine should be avoided in older adults (American Geriatrics Society Beers Criteria® Update Expert Panel, 2012, 2019). There are several safer alternative therapeutic options for older adults, such as citalopram, escitalopram, sertraline, venlafaxine, mirtazapine, and bupropion (Wiese, 2011).

The purpose of this study was to examine patterns in paroxetine prescription rates throughout the United States among Medicare patients. This study has important implications for education and policy reform regarding paroxetine use in older adults.

## Methods

### Procedures

Annual (2015-2020) paroxetine prescription rates and costs were obtained for Medicare Part D patients. We evaluated the Medicare Specialty Utilization and Payment Data for paroxetine prescription rates per state (Centers for Medicare & Medicaid Services, n.d.-b). Prescription rates were reported per thousand Medicare Part D enrollees. Number of specialties per specialty was obtained from Medicare Physician and Other Practitioners by Specialty and Service (Centers for Medicare & Medicaid Services, n.d.-c). Procedures were approved as exempt by the Geisinger IRB.

### Data Analysis

Patterns in the number of national prescriptions of generic, brand, and their sum were compared for paroxetine. One-sample z-tests were conducted to determine if any states’ annual prescription rates were significantly different from the state average for that respective year. The ratio of total Medicare spending for generic versus brand was also calculated.

To understand specialty-type variations, ratios were calculated. Percent of paroxetine prescriptions from a particular specialty relative to all specialties was calculated. The aforesaid percent was divided by the percent of providers in Medicare who belong to a particular specialty. Ratios greater than 1.0 suggested the specialty was overrepresented, and ratios under 1.0 suggested the specialty was underrepresented for paroxetine prescriptions. Specialties for whom taxonomy codes could not be mapped to a Medicare specialty code were excluded. Data was analyzed using Excel and figures illustrating the ten specialties with the highest ratios per year were constructed using GraphPad Prism.

## Results

Figure 2 shows that population-corrected paroxetine use and spending gradually decreased across from 2015 to 2020. There were 122.61 prescriptions per thousand Medicare enrollees in 2015. This decreased by 34.52% to 116.80 prescriptions per thousand Medicare enrollees in 2020. There was $90.88 million dollars spent in 2015. This value decreased by 16.69% to $75.71 million dollars in 2020. Generic paroxetine consistently constituted ≥ 99.2% of all prescriptions, and increased by 0.5%, when spending on generics consistently constituted around 90% of all paroxetine spending.

**Figure 2.**
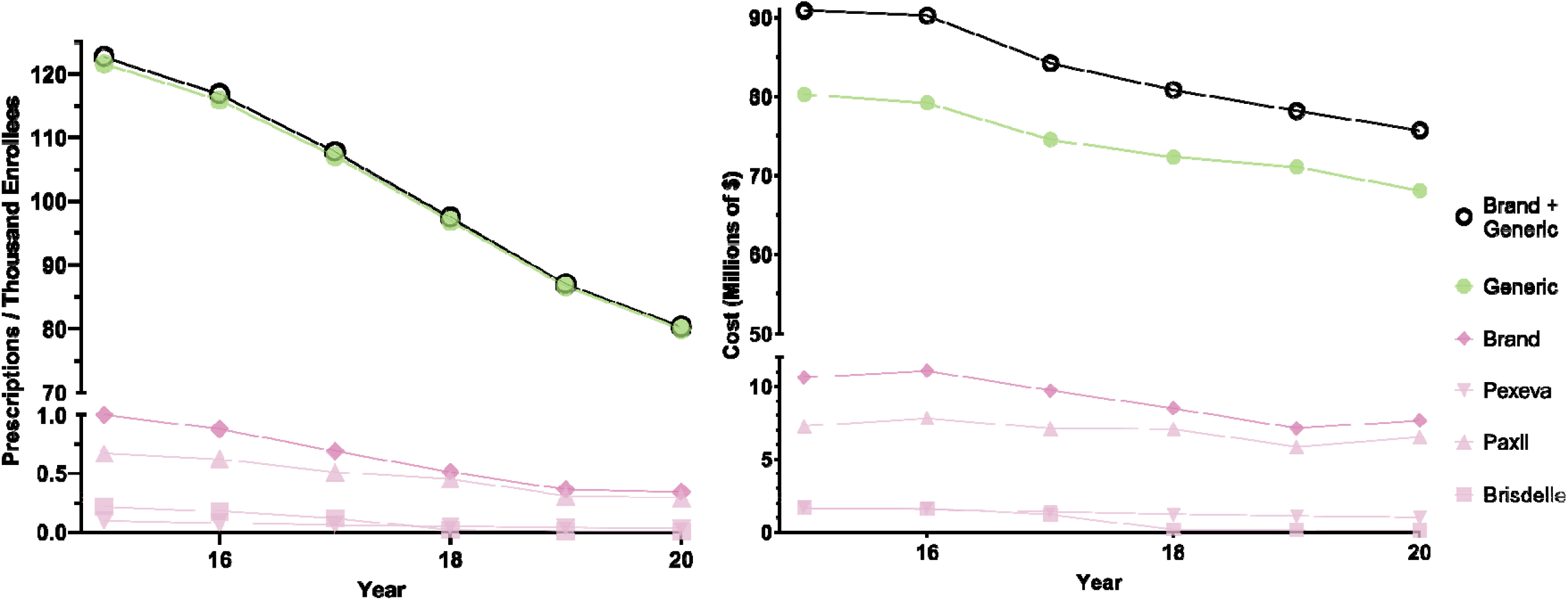
National Medicare prescription rates (left) and Medicare spending (right) for paroxetine for 2015-2020.

Figure 3 and Supplemental Figures 1-5 show wide state-level variation in 2015 (4.6 fold), 2016 (5.2 fold), 2017 (5.3 fold), 2018 (5.2 fold), 2019 (5.2 fold), and 2020 (5.0 fold). Kentucky was always the highest prescribing state with a significantly greater number of prescriptions than the mean number of state prescriptions in all years examined. North Dakota was the second highest prescribing state, except for 2015 and 2017, and had a significantly higher number of paroxetine prescriptions than the mean number of state prescriptions in 2016, 2018, and 2019. Alaska had significantly more paroxetine prescriptions than average in 2015. On the other end of the spectrum, Hawaii, the lowest prescribing state, had a significantly lower number of prescriptions than the mean number of state prescriptions in all years examined. The District of Columbia, consistently the second or third lowest prescribing municipality, prescribed significantly less paroxetine than other states in 2015, 2016, and 2018.

**Figure 3.**
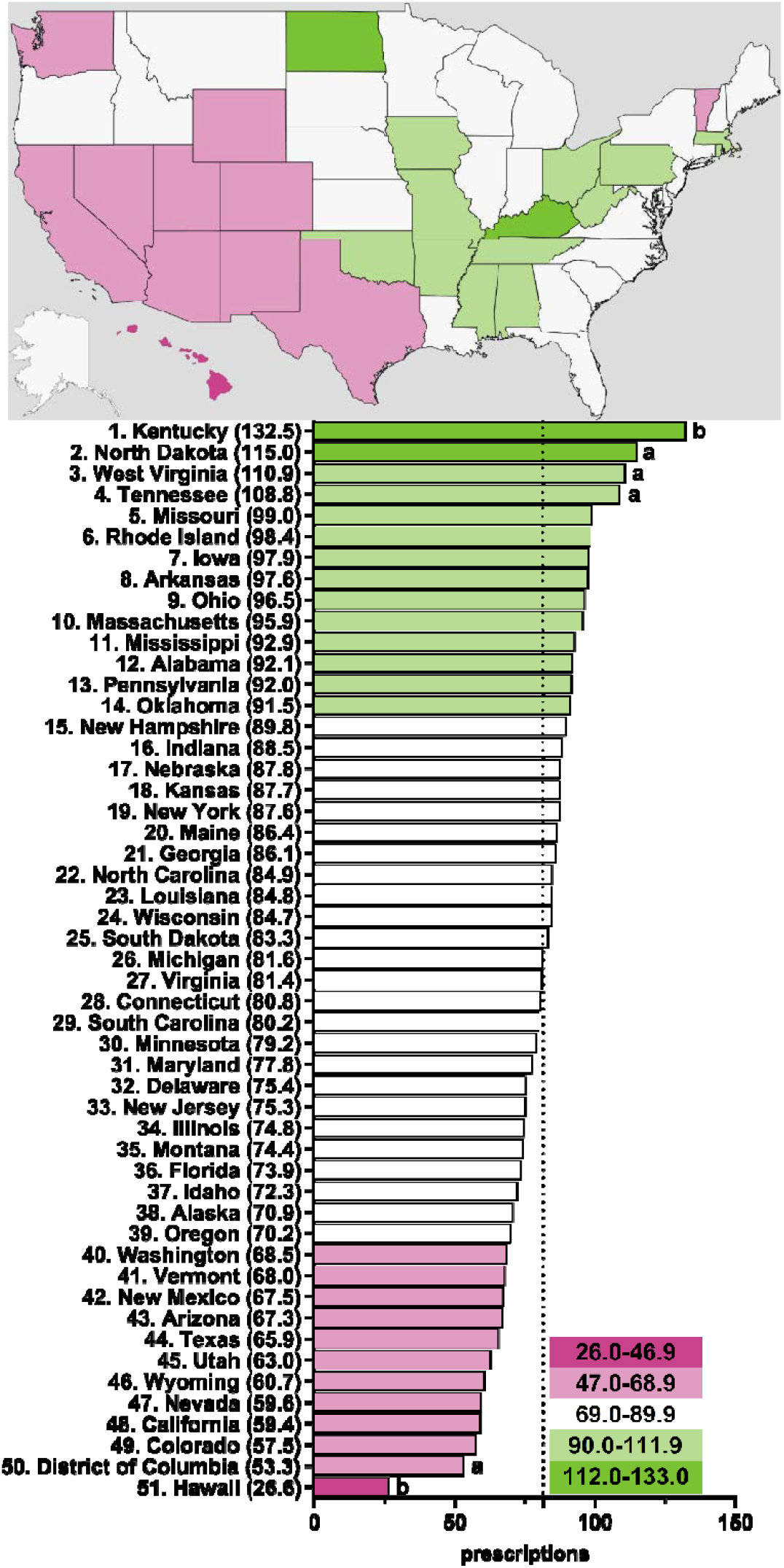
Paroxetine prescriptions per thousand Medicare Part D enrollees heatmap (top) and population-corrected prescription rate per state (bottom) in 2020. ^a^ indicates ≥1.50 SD (17.4) from the mean (81.5), denoted by the dotted line. ^b^ indicates ≥1.96 SD from the mean.

Figure 4 and Supplemental Figure 2 shows the specialties with the highest ratios of percent of paroxetine prescriptions to percent of practitioners in Medicare who belong to that respective specialty. For all years examined, neuropsychiatry, geriatric medicine, geriatric psychiatry, psychiatry, family practice, internal medicine, general practice, and certified clinical nurse specialists had ratios above 1, with neuropsychiatry and geriatric medicine consistently having the highest ratios, except for geriatric psychiatry having a higher ratio than geriatric medicine in 2020. Neuropsychiatry and psychiatry notably and consistently had the highest ratios for brand paroxetine prescriptions for all years examined.

**Figure 4.**
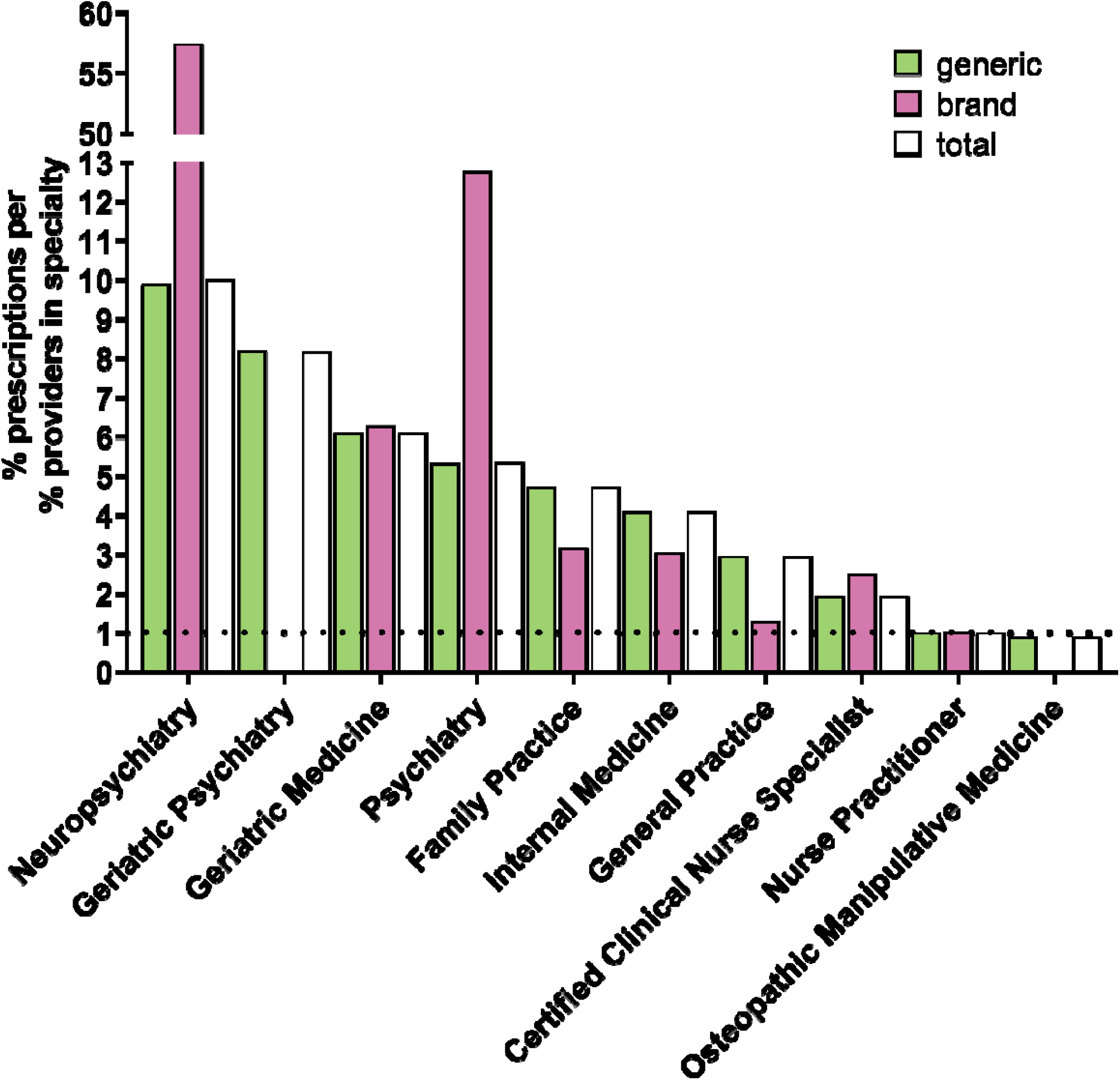
*Specialty types that prescribe the most paroxetine to Medicare Part D enrollees for 2020*. Specialty types that had the highest ratio of percent of paroxetine prescriptions to percent of providers in Medicare who belong to that respective specialty. Dotted line denotes ratio of 1.0.

## Discussion

Despite the identification in 2012 of paroxetine as a PIM in older adults (American Geriatrics Society Beers Criteria® Update Expert Panel, 2012), we found appreciable use (123 prescriptions / 1,000 enrollees) among older (≥65) US Medicare patients in 2015. It is encouraging that the data in this study show that paroxetine use in a population who is not generally recommended has been in a steady decline for the generic formulations. Similarly, branded formulations, although uncommon (Buttorff et al., 2020), also declined, except for a mild relative increase in Paxil between 2019 and 2020. Continued recommendations against the use of paroxetine among older adults (AGS, 2019), as well as increased therapeutic alternatives likely has contributed to the decline so far observed and serves as potential means for the decline to continue (Marks et al., 2008; Nevels et al., 2016; Wiese, 2011). Previously mixed recommendations (American Geriatrics Society Beers Criteria® Update Expert Panel, 2012) on the use of paroxetine in older adults may be contributing to its abatement among older adults in recent years. It is worth noting that there are individual considerations that may make the evidence against paroxetine not applicable to a given provider-patient relationship.

Kentucky and North Dakota are the states that appear to have the largest potential benefit to the introduction and/or improvement of education and/or policy related to the recommended uses of paroxetine. Similarly, the specialties that have the most potential benefit from education and policy changes related to paroxetine use among older adults are: neuropsychiatry, geriatric medicine, geriatric psychiatry, psychiatry, family practice, internal medicine, general practice, and certified clinical nurse specialists. More research needs to be done to understand the geographic and specialty variation seen in this study to draw conclusions as to potential causes for the unexpected heterogeneity.

Although the Medicare program serves 94% of non-institutionalized persons age 65 and older, some limitations of this study are noteworthy (Administration on Aging, 2021). First, one-sixth of those who have Medicare are under the age of 65, and one-quarter of people with Medicare do not have Part D coverage (Centers for Medicare & Medicaid Services, n.d.-a). Second, further study with other databases will be necessary to characterize whether paroxetine was prescribed for anxiety disorders, major depression, or an off-label indication (e.g., sleep disturbance or sedation in nursing home residents). Third, it is worth noting despite significant evidence suggesting paroxetine has significantly higher risk when used in the older adult population, some studies have demonstrated no important cognitive adverse effects in this population (Cassano et al., 2002; Nebes et al., 1999).

Regardless, the persistent relative frequent use of paroxetine in this population is concerning, especially given the existence of safer alternatives (American Geriatrics Society Beers Criteria® Update Expert Panel, 2012, 2019; Wiese, 2011). Even more, though the spending on paroxetine by older adults illustrated in Figure 2 averages around a sizable $80 million per year, it does not capture the inpatient expenditures and treatments that older adults may need as consequence of inappropriate use of paroxetine, which impacts the higher spending in high-risk populations more than pharmaceutical costs (Pritchard et al., 2016). With the continued prevalent use of paroxetine in older adults, implementation of ongoing computerized reminder systems may be worth consideration (Soumerai et al., 2005). Moreover, the overall high frequency of adverse drug events (Beijer & de Blaey, 2002; M. Chan et al., 2001; Page & Ruscin, 2006) and use of PIMs (Insani et al., 2021) in older adults suggests that routine review of this population’s medication regimen with a standardized tool, such as Beers (American Geriatrics Society Beers Criteria® Update Expert Panel, 2012, 2019), STOPP (Hamilton et al., 2011), or START (Barry et al., 2007), would likely have a substantial impact on improving the healthcare outcomes of the age demographic that uses most health care resources (Pretorius et al., 2013).

The observed decline in paroxetine utilization and expenditure among older adults mirrors decreasing usage of paroxetine in other populations, likely due to the advent of newer, more efficacious and tolerable antidepressants. Thus, the decreasing pattern in paroxetine use found in this study may merely reflect the general decrease in use of paroxetine, not due to recommendations to avoid paroxetine use in older adults. For example, in a study on antidepressant use and expenditure in six major cities in China that found increased usage of SSRIs, the percentage of the total expenditure accounting for paroxetine, significantly decreased from 21.9% in 2013 to 11.9% in 2018 (Yu et al., 2020). In a study on antidepressant use in France which found increased use of SSRIs, the share of paroxetine halved from 14.1% in 2009 to 6.7% in 2016 in the overall population and from 15.4% in 2009 to 7.0% in 2016 in 12- to 17-year-olds (Revet et al., 2018). Future studies could explore if the decrease in paroxetine use observed in this study is unique to older adults or if it is consistent across other age demographics.

Though the focus of this study was understanding paroxetine use among older adults, there are many other medications and drug interactions that contribute to serious adverse events in older adults. Future directions may include characterizing the pharmacoepidemiology of these other high-risk medications in older adults, as well as monitoring polypharmacy among older adults with paroxetine and other drugs. Additionally, examining how the prescribing patterns of these high-risk medications has changed as result of the implementation of different clinical tools, such as the Beers, STOPP, and START criteria would be meaningful.

## Supporting information

Supplemental- Analyzed Data 1

Supplemental- Analyzed Data 2

## Data Availability

All data produced in the present study are available upon reasonable request to the authors and available as supplemental material on medrxiv.org

https://data.cms.gov/provider-summary-by-type-of-service/medicare-part-d-prescribers

## Acknowledgements

BJP was supported by HRSA (D34HP31025). Software used for this research was provided by NIEHS (T32-ES007060-31A1).

## Notes

**Disclosures:** BJP’s research is supported by the Pennsylvania Academic Clinical Research Center and the Health Resources and Services Administration (D34HP31025). Prior (2019 – 2021) osteoarthritis research was supported by Pfizer and Eli Lilly. The other authors have no disclosures.

### Competing Interest Statement

Disclosures: Research of BJP is supported by the Pennsylvania Academic Clinical Research Center and the Health Resources and Services Administration (D34HP31025). Prior (2019 to 2021) osteoarthritis research was supported by Pfizer and Eli Lilly. The other authors have no disclosures.

