## Supplemental Figures for "Specialty-type and state-level variation in paroxetine use among older adult patients"

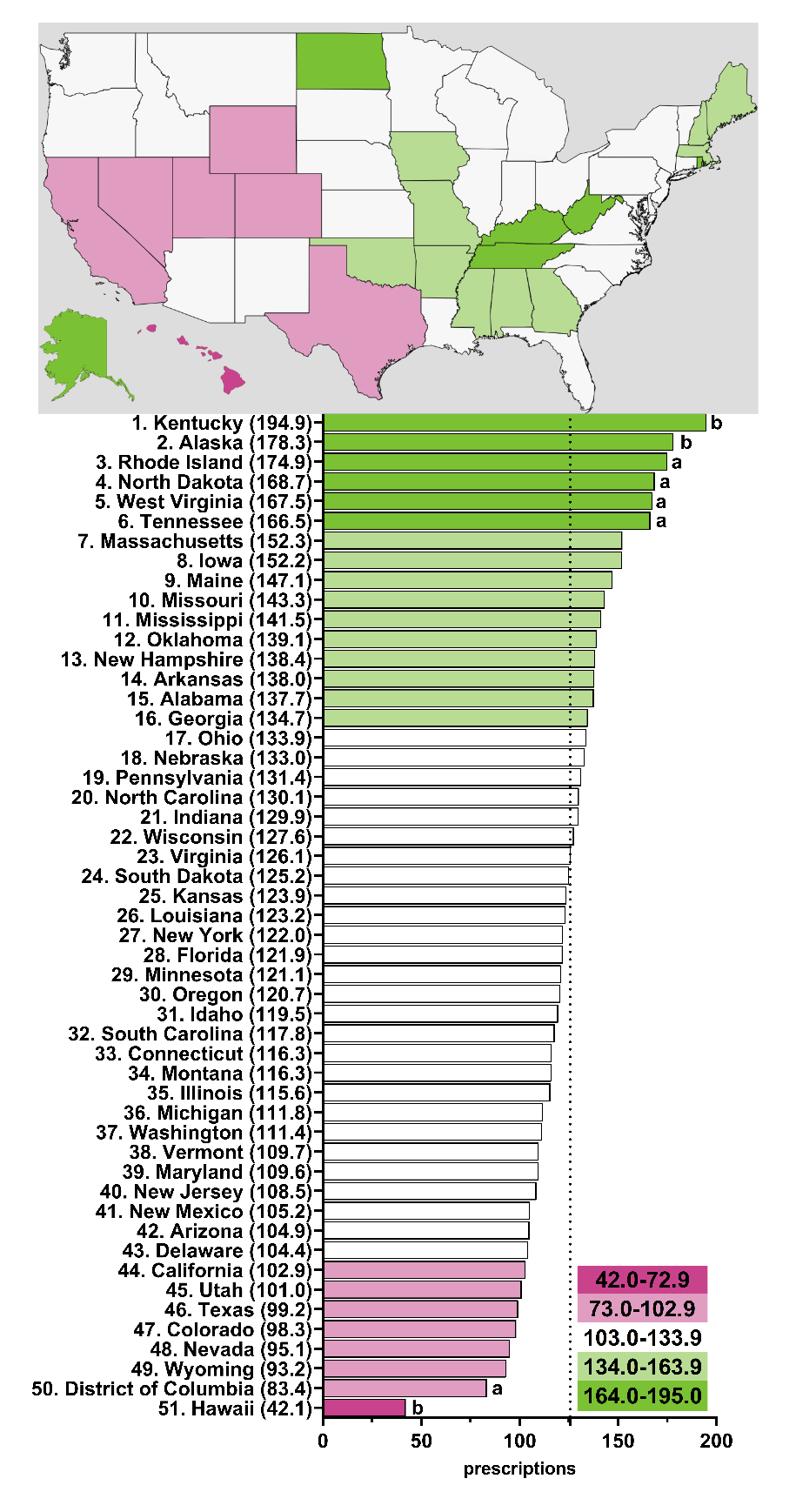

**Supplemental Figure 1.** Paroxetine prescriptions per thousand Medicare Part D enrollees heatmap (top) and population-corrected prescription rate per state (bottom) in 2015. ^a^ indicates ≥1.50 SD (26.2) from the mean (125.7), denoted by the dotted line. ^b^ indicates ≥1.96 SD from the mean.

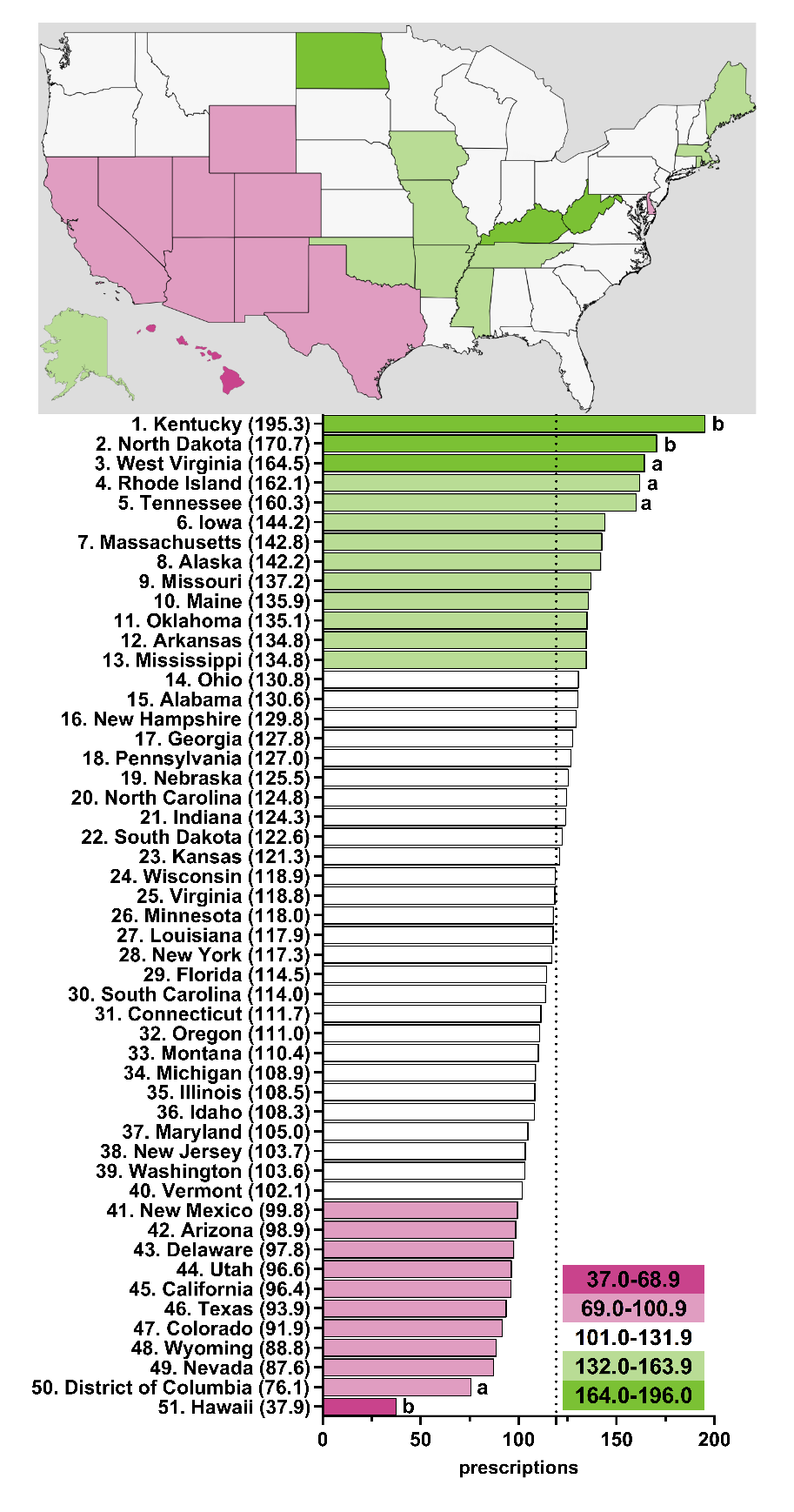

**Supplemental Figure 2.** Paroxetine prescriptions per thousand Medicare Part D enrollees heatmap (top) and population-corrected prescription rate per state (bottom) in 2016. ^a^ indicates ≥1.50 SD (25.6) from the mean (119.3), denoted by the dotted line. ^b^ indicates ≥1.96 SD from the mean.

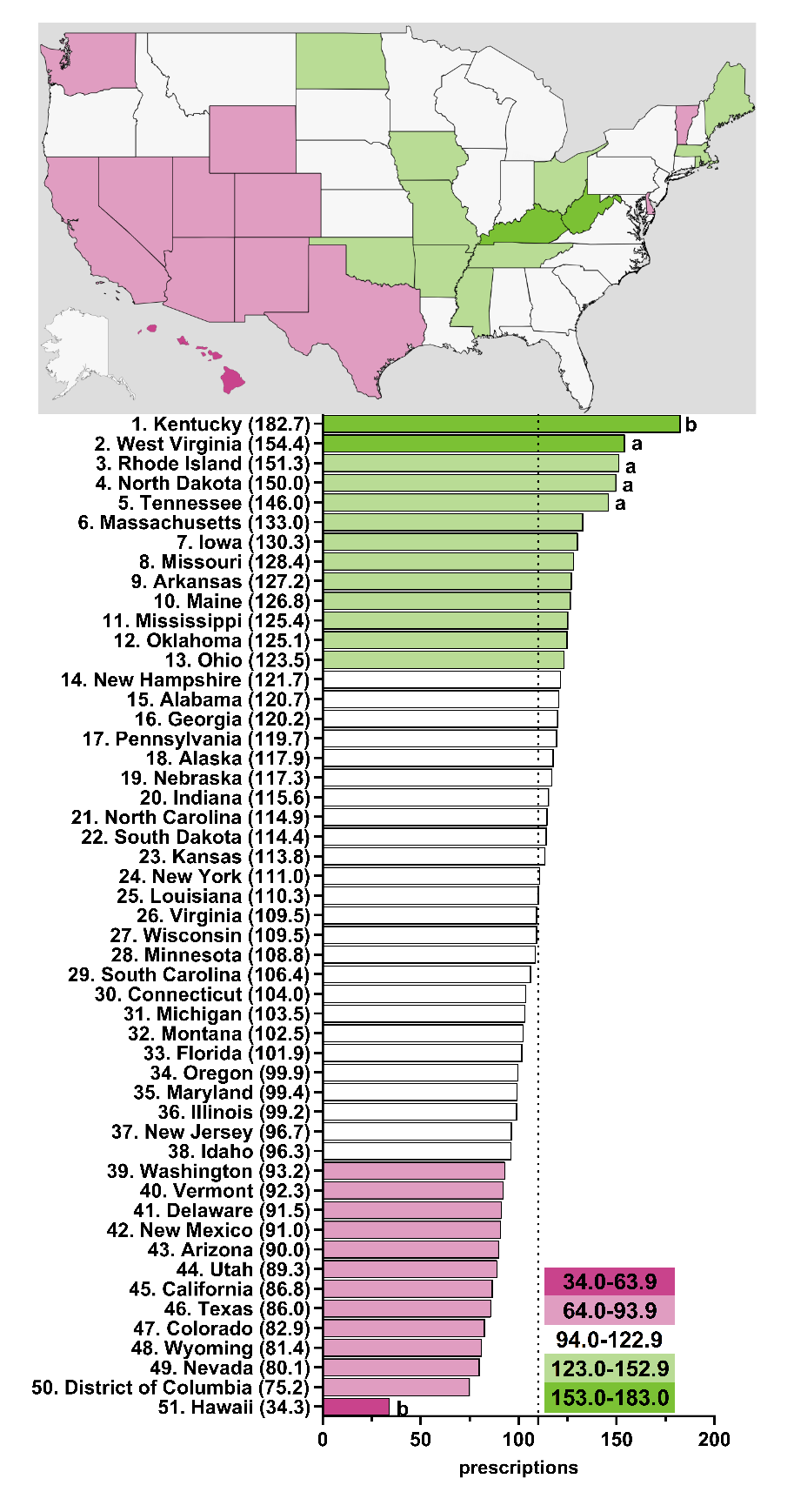

**Supplemental Figure 3.** Paroxetine prescriptions per thousand Medicare Part D enrollees heatmap (top) and population-corrected prescription rate per state (bottom) in 2017. ^a^ indicates ≥1.50 SD (23.7) from the mean (110.1), denoted by the dotted line. ^b^ indicates ≥1.96 SD from the mean.

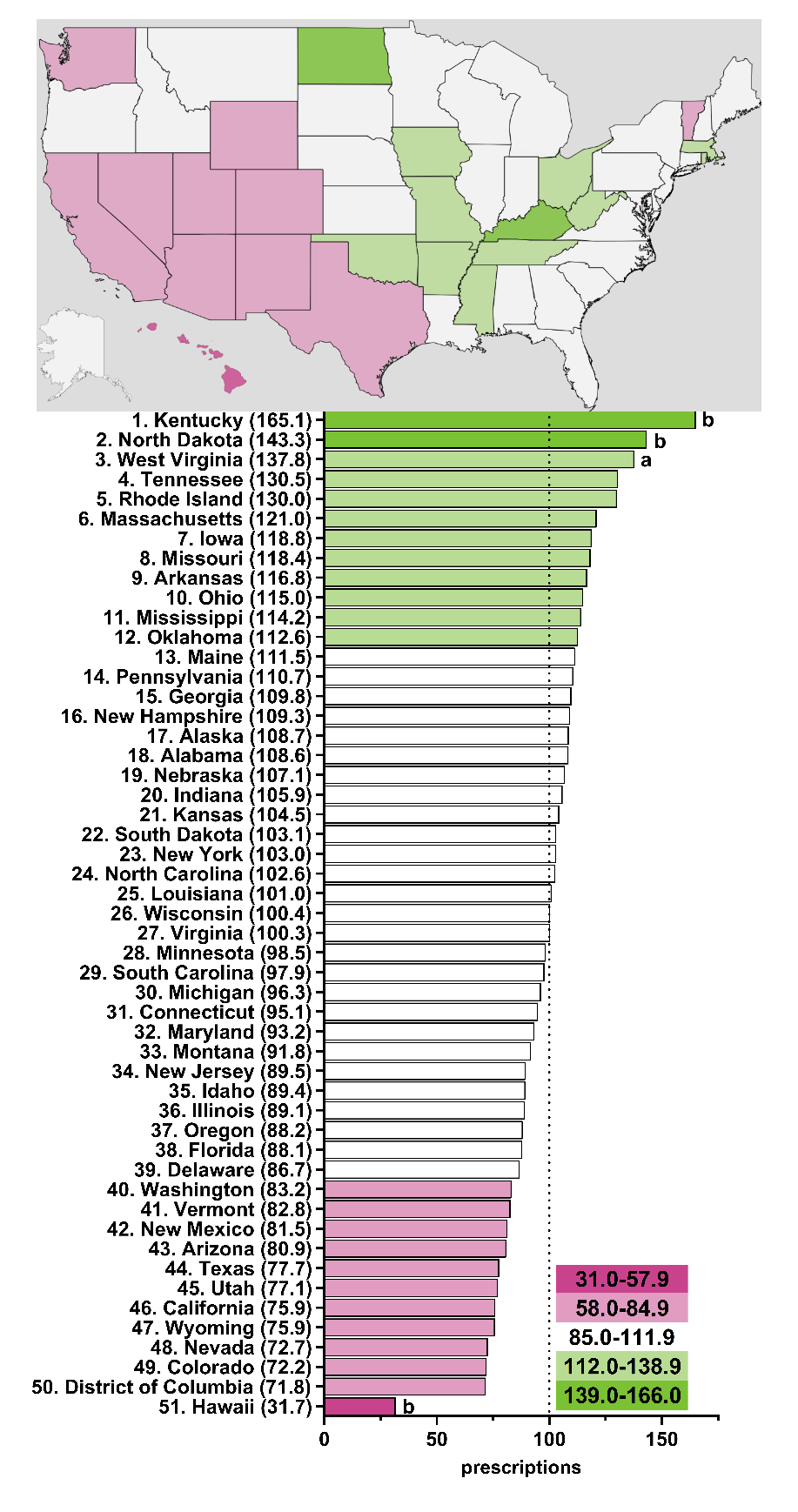

**Supplemental Figure 4.** Paroxetine prescriptions per thousand Medicare Part D enrollees heatmap (top) and population-corrected prescription rate per state (bottom) in 2018. ^a^ indicates ≥1.50 SD (21.5) from the mean (99.9), denoted by the dotted line. ^b^ indicates ≥1.96 SD from the mean.

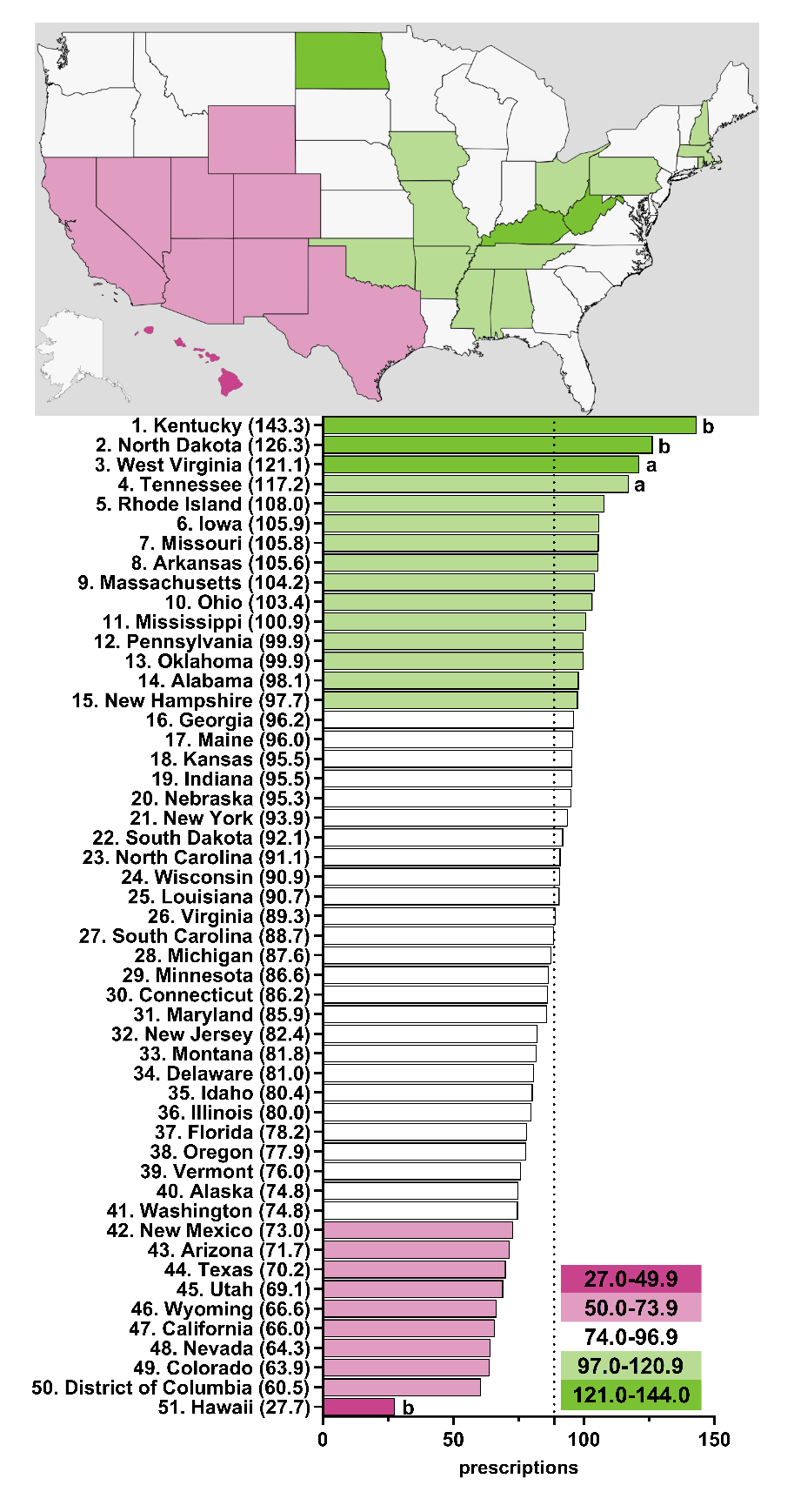

**Supplemental Figure 5.** Paroxetine prescriptions per thousand Medicare Part D enrollees heatmap (top) and population-corrected prescription rate per state (bottom) in 2019. ^a^ indicates ≥1.50 SD (18.7) from the mean (88.6), denoted by the dotted line. ^b^ indicates ≥1.96 SD from the mean.

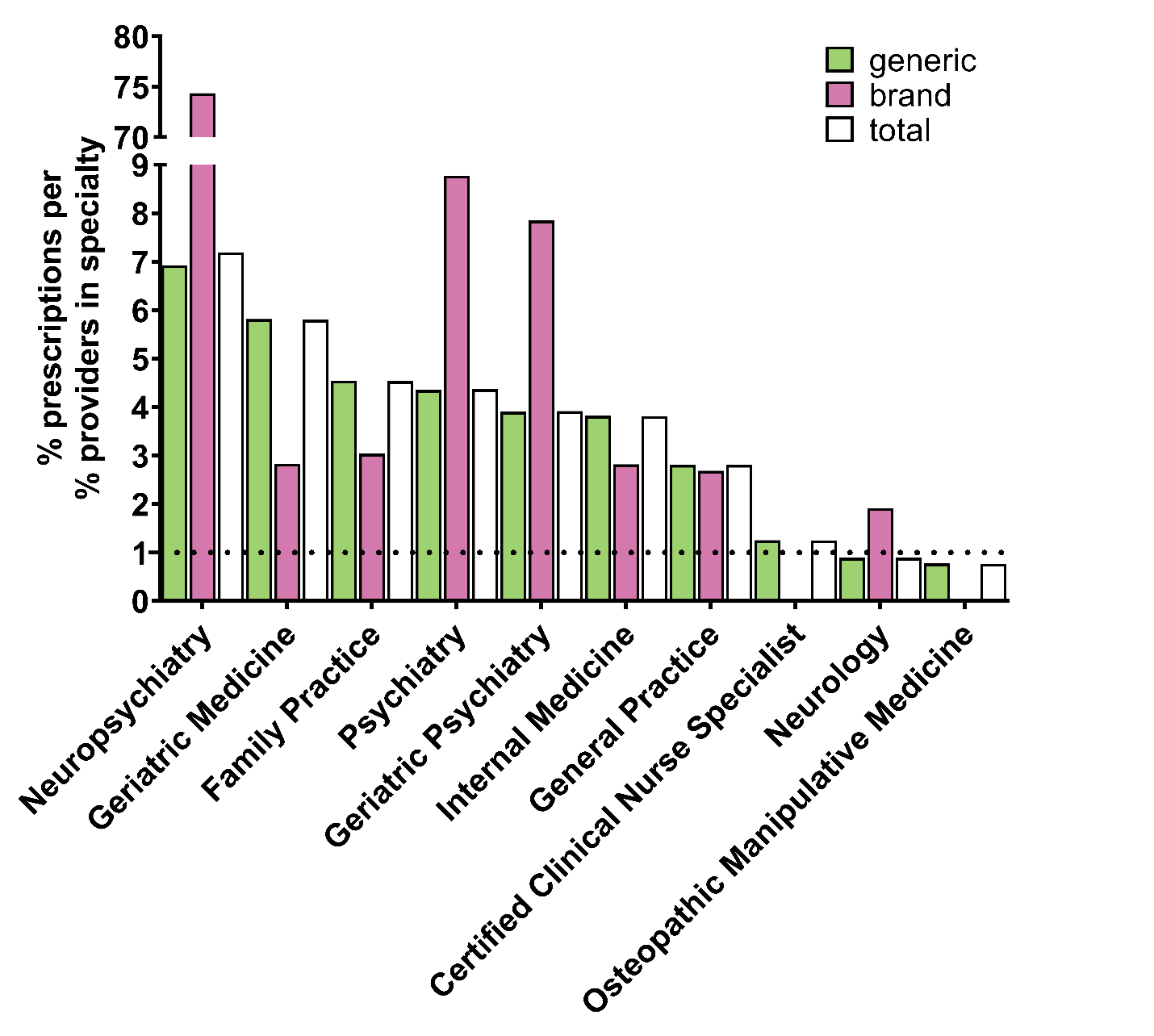

**Supplemental Figure 6.** *Specialty types that prescribe the most paroxetine to Medicare Part D enrollees for 2015.* Specialty types that had the highest ratio of percent of paroxetine prescriptions to percent of providers in Medicare who belong to that respective specialty. Dotted line denotes ratio of 1.0.

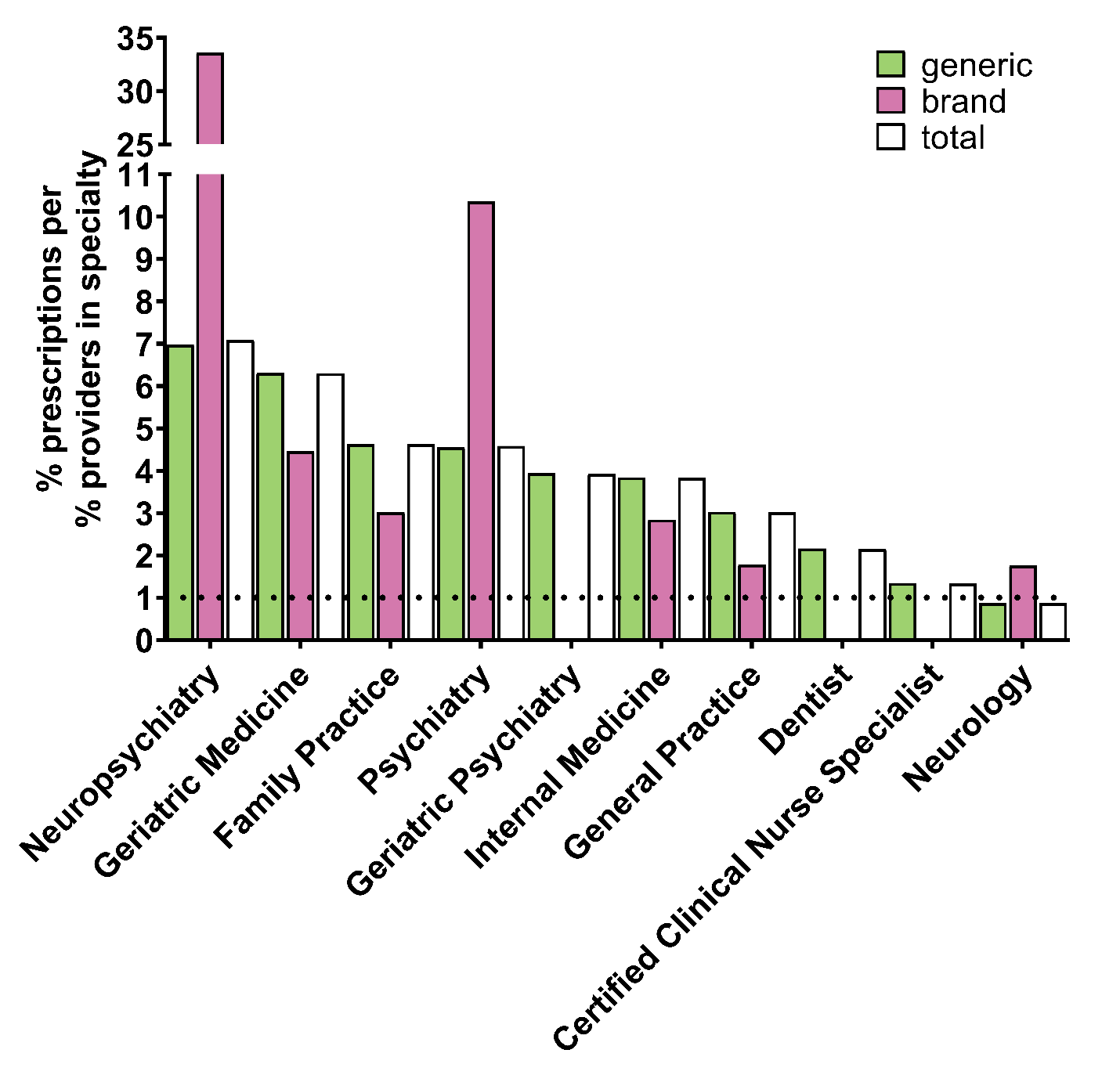

**Supplemental Figure 7.** *Specialty types that prescribe the most paroxetine to Medicare Part D enrollees for 2016.* Specialty types that had the highest ratio of percent of paroxetine prescriptions to percent of providers in Medicare who belong to that respective specialty. Dotted line denotes ratio of 1.0.

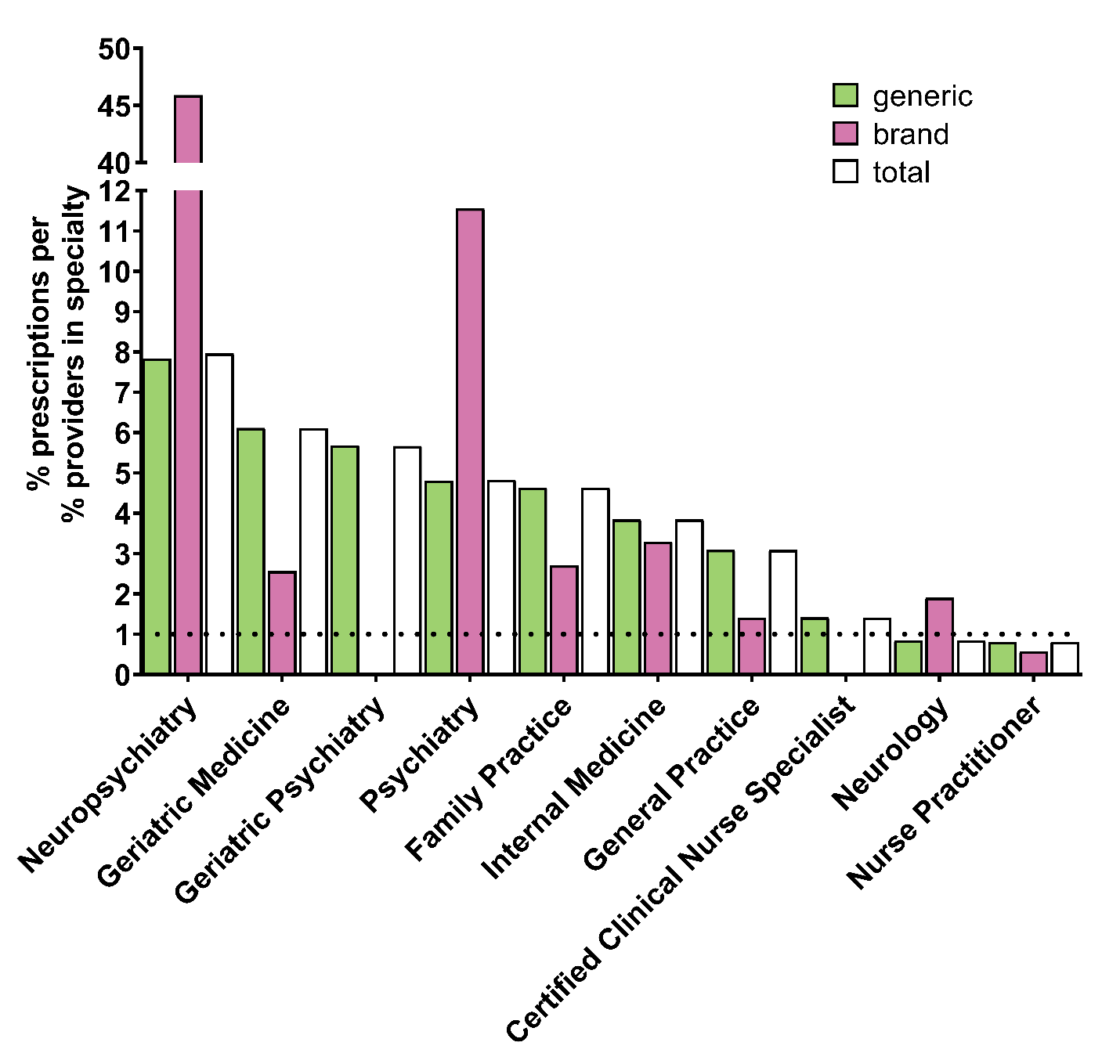

**Supplemental Figure 8.** *Specialty types that prescribe the most paroxetine to Medicare Part D enrollees for 2017.* Specialty types that had the highest ratio of percent of paroxetine prescriptions to percent of providers in Medicare who belong to that respective specialty. Dotted line denotes ratio of 1.0.

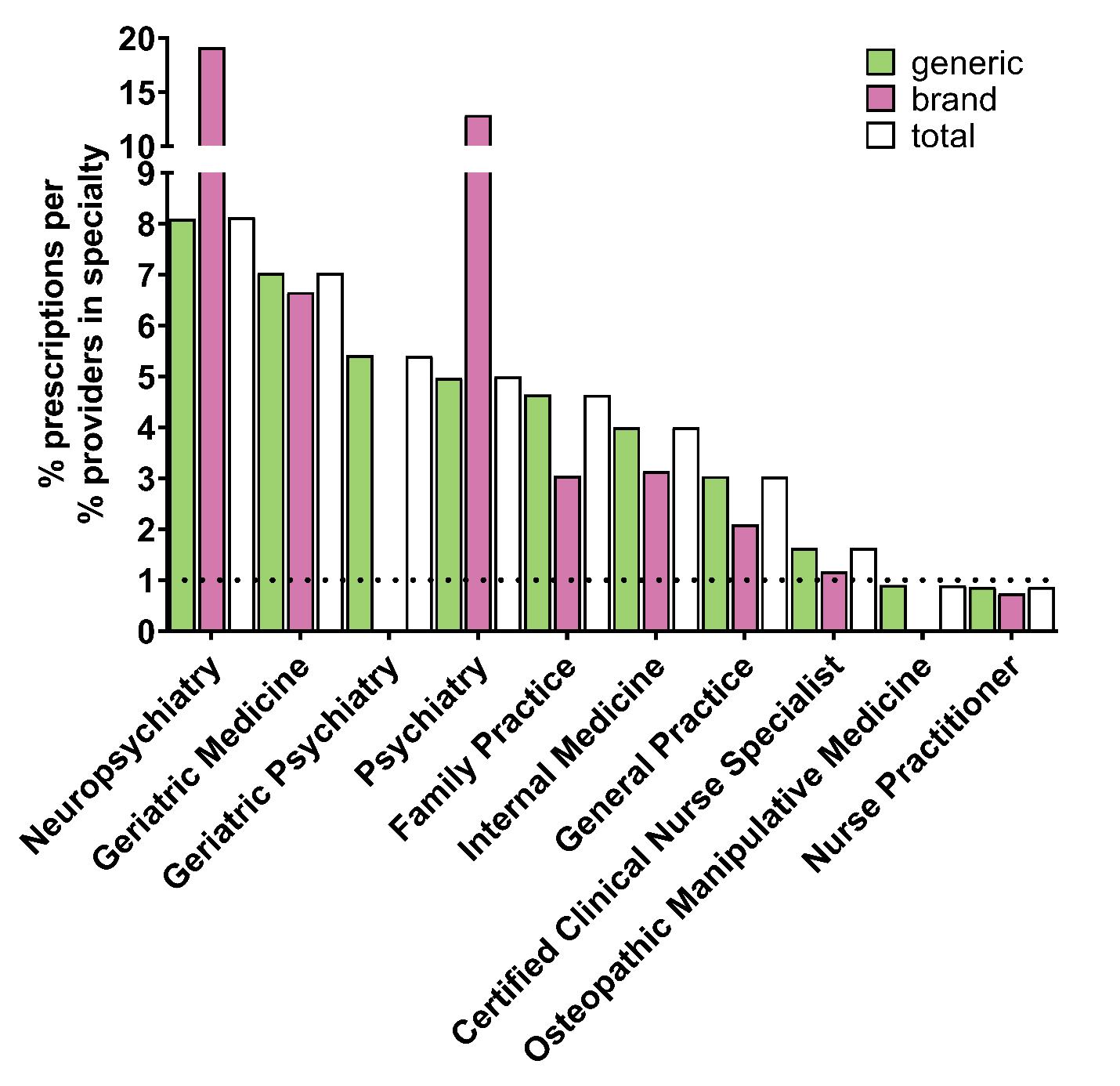

**Supplemental Figure 9.** *Specialty types that prescribe the most paroxetine to Medicare Part D enrollees for 2018.* Specialty types that had the highest ratio of percent of paroxetine prescriptions to percent of providers in Medicare who belong to that respective specialty. Dotted line denotes ratio of 1.0.

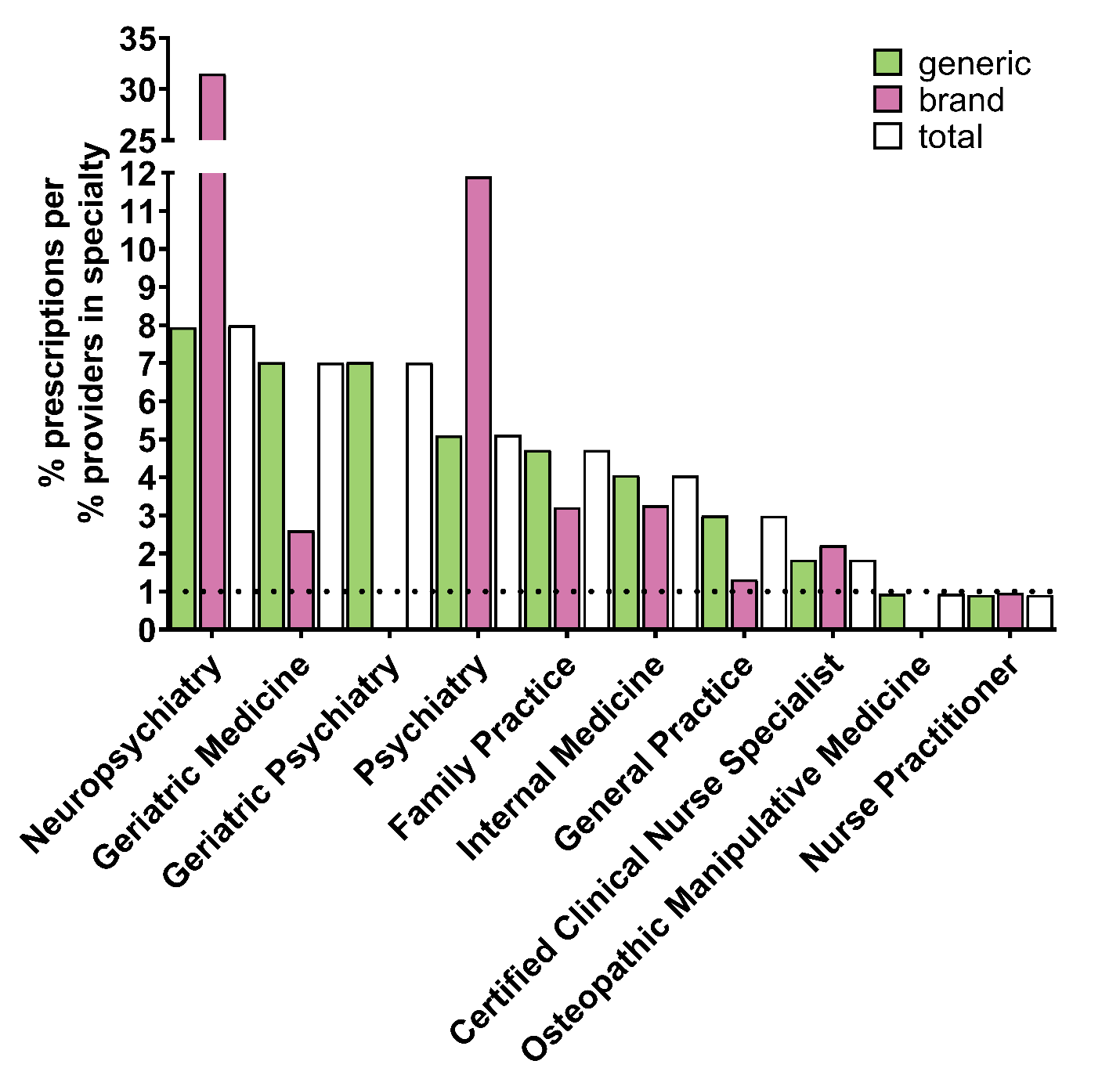

**Supplemental Figure 10.** *Specialty types that prescribe the most paroxetine to Medicare Part D enrollees for 2019.* Specialty types that had the highest ratio of percent of paroxetine prescriptions to percent of providers in Medicare who belong to that respective specialty. Dotted line denotes ratio of 1.0.
